# Temporal Relationship of Computed and Structured Diagnoses in Electronic Health Record Data

**DOI:** 10.1101/2019.12.28.19015628

**Authors:** Wade L. Schulz, H. Patrick Young, Andreas Coppi, Bobak J. Mortazavi, Zhenqiu Lin, Raymond A. Jean, Harlan M. Krumholz

## Abstract

The electronic health record (EHR) holds the prospect of providing more complete and timely access to clinical information for studies, quality assessments, and quality improvement compared to other data sources, such as administrative claims. Our goal was to assess the completeness and timeliness of structured diagnoses in the EHR compared to computed diagnoses for hypertension (HTN), hyperlipidemia (HLD), and diabetes mellitus (DM). We determined the amount of time for a structured diagnosis to be recorded in the EHR from when an equivalent diagnosis could be computed from other structured data elements, such as vital signs and laboratory results. Using our local instance of EHR data in the PCORnet common data model (CDM) with encounters from January 1, 2012 through February 10, 2019, we identified patients with at least two observations above threshold separated by at least 30 days. The thresholds were outpatient blood pressure of ≥ 140/90 mmHg, any low-density lipoprotein ≥ 130 mg/dl, or any hemoglobin A1c ≥ 7%, respectively. The primary measure was the length of time between the computed diagnosis and the time at which a structured diagnosis could be identified within the EHR history or problem list. We found that 39.8% of those with HTN, 21.6% with HLD, and 1.0% with DM did not receive a corresponding structured diagnosis recorded in the EHR. For those who received a structured diagnosis, a mean of 389, 198, and 106 days elapsed before the patient had the corresponding diagnosis of HTN, HLD, or DM, respectively, recorded in the EHR. We identified a marked temporal delay between when a diagnosis can be computed or inferred and when an equivalent structured diagnosis is recorded within the EHR. These findings demonstrate the continued need for additional study of the EHR to avoid bias when using observational data and reinforce the need for computational approaches to identify clinical phenotypes.

## Introduction

Despite the rapid digitization of healthcare, the current research enterprise remains inefficient. Randomized control trials (RCTs), which remain the gold standard, are costly, time-consuming, and capture only a small cross-section of patients, which limits their generalizability.^1,2^ To accelerate the pace of discovery, investigators and regulatory agencies have increasingly focused on real-world data (RWD), defined as data collected outside of a traditional research environment, as a source of information.^3–5^ Real-world data sources include administrative claims and discharge databases, clinical registries, and electronic health records (EHRs), among others. Despite increased access to these digital data, it remains important for investigators to be aware of and account for limitations in these observational data sets during study design and analysis.^6–9^

The creation of patient cohorts and study endpoints often requires the identification of clinical diagnoses. These populations and endpoints are frequently characterized by diagnostic codes, such as International Statistical Classification of Diseases and Related Health Problems 10^th^ Revision Clinical Modification (ICD-10-CM) codes.^10^ Observational and outcomes research have long used administrative claims and registries as a source of this information.^11–18^ However, these repositories come with the known limitations of significant time delays in availability and a lack of detailed clinical records.^19,20^ In addition, significant costs are associated with manual abstraction for disease-specific registries.^21^ Because of these limitations and the increased access to detailed EHR data, investigators have increasingly focused on the EHR to provide the data needed to support a wide range of studies.

Information obtained from the EHR has the potential to provide near real-time access to a more complete data set than can be provided from other RWD sources.^19,20^ The EHR is the primary repository of a patient’s clinical history, but the data elements needed to represent a patient’s history can be found in many locations, from structured fields in the history and problem list to unstructured clinical notes.^9^ Prior work has shown that patient history and problem lists within the EHR can be incomplete and contain frequent errors.^22–25^ Even for a relatively straightforward diagnosis such as hypertension, researchers from the OneFlorida clinical data research network found that as many as 30% of those they identified with hypertension by means of clinical measurements recorded in the EHR were missing the associated structured diagnostic code,^26^ similar to results found by an earlier study from Stanford.^27^

Yet studies based on EHR data frequently use, sometimes solely, structured diagnostic codes to create cohorts and identify outcomes.^28,29^ Since administrative claims are ultimately derived from these structured fields, the EHR is likely not significantly worse than claims-based sources.

Investigators have also demonstrated that structured diagnostic codes can provide valuable information when analyzed in the appropriate context.^30,31^ Therefore, while limitations to the use of structured diagnoses from the EHR exist, they remain frequently used and ongoing study can increase the value of results generated from these sources.

A primary goal of EHR-based studies is access to near real-time information,^32,33^ but it remains an indirect assessment of a patient’s state due to how the EHR is used in clinical workflows.^34^ While data may be extracted immediately, clinical workflows and decision making may impact the timeliness of data entry into the EHR, particularly within structured data elements. Because of this, the extent to which structured diagnoses may be missing at the time of analysis may be greatly underestimated and, therefore, next generation phenotypes may be able to provide more accurate and timely information.

In this study, we determine how structured diagnostic codes in the patient history and problem list compare to computed diagnoses for hypertension (HTN), hyperlipidemia (HLD), and diabetes mellitus (DM). We selected these three diagnoses because they can be efficiently computed from structured clinical and laboratory data. We extend on prior work in the field to identify not only the completeness of structured diagnostic codes, but also the temporal association between the computed diagnosis and manual recording of an equivalent diagnostic code within the clinical record.

## Methods

### Data Sources

We created our data set from a complete extract of the Yale New Haven Health clinical data warehouse (Epic Caboodle) that was transformed into the PCORnet Common Data Model (CDM) on February 11, 2019 using our local data analytics platform.^35^ The Caboodle source tables and supporting terminology tables were transformed into the *demographic, encounter, diagnosis, condition, lab_result_cm*, and *vital* PCORnet CDM tables, which were used for analysis. Diagnosis and condition source were categorized as defined in the PCORnet CDM.^36^ As a data quality study based on existing and deidentified data, this work was not classified as human subjects research and did not require Institutional Review Board approval.

### Phenotype Definitions

We collected information to assess HTN, HLD, and DM. We excluded data with dates earlier than January 1, 2012 as these represented a sparse fraction of the raw data with unreliable dates of onset due to their transfer between prior EHR systems. The most recent date of measurement or sample collection included in the analysis was restricted to on or before August 11, 2018, whereas diagnosis events were current up to February 10, 2019, allowing a period of at least 26 weeks (approximately six months) between the most recent measurement or result and the final date available. If multiple measurements were performed on the same day, then the minimum value of the measurement was selected to provide the most restrictive threshold. Only outpatient blood pressure readings, as annotated within the PCORnet CDM, were used, but laboratory results were extracted from all encounter settings. Relevant laboratory results for LDL and A1c were identified by LOINC codes (LDL: ‘13457-7’,’18262-6’; A1c: ‘4548-4’) code or internal EHR codes (various and specific to our institution).

We flagged measurements as a ‘signal of disease’ whenever they exceed a specific threshold. For HTN, a measurement was flagged as a ‘signal of disease’ if either the minimum systolic reading was ≥ 140 mmHg or the minimum diastolic reading was ≥ 90 mmHg. For HLD, the threshold was an LDL ≥ 130 mg/dL and for DM the threshold was an A1c ≥ 7%. For all three conditions, if any two measurements taken at least 30 days apart were found above the threshold (abnormal is high in all three cases), the patient was considered to have a computed diagnosis of disease on the date of the second signal.

For each condition and diagnosis code system, the first 3 or 5 characters of the code string were matched against ICD-10-CM and ICD-9-CM parent codes (Table 1). Patients with a diagnosis present prior to the first signal were flagged as having an existing diagnosis. For patients without a prior diagnosis or computed diagnosis as defined above, the first date a matching diagnosis code (Table 1) was recorded in the EHR, if present, was logged along with its origin (provider-entered billing diagnosis; provider-entered encounter diagnosis or problem list entry; or diagnosis code from returned claims) and the date of the patient’s most recent encounter with the health system. A final data set consisting of the computed signal dates, computed and structured diagnosis dates, date of most recent encounter, and diagnosis origin was used for analysis.

**Table 1:** Diagnosis criteria and ICD-9-CM and ICD-10-CM code groups that were used for each condition to determine computed signals of disease and recorded diagnoses. Abbreviations: *ICD-9-CM, International Classification of Diseases, Ninth Revision, Clinical Modification; ICD-10-CM, International Classification of Diseases, Tenth Revision, Clinical Modification; LDL, Low-density lipoprotein cholesterol*

| Condition | Signal and Diagnosis Criteria |
| --- | --- |
| **Hypertension** | **Denominator:** Outpatient systolic $\geq 140$ or diastolic $\geq 90$ (mmHg).<br>2 readings $\geq 30$ days apart and no prior diagnosis.<br>**Numerator:** New diagnosis of any of the following:<br>ICD-9 groups: 401, 402, 403, 404, 405, 642<br>ICD-10 groups: I10, I11, I12, I13, I15, I16, I27, O13 |
| **Diabetes** | **Denominator:** A1c $\geq 7$ (%).<br>2 readings $\geq 30$ days apart and no prior diagnosis.<br>**Numerator:** New diagnosis of any of the following:<br>ICD-9 groups: 249, 250<br>ICD-10 groups: E08, E09, E10, E11, E13 |
| **Hyperlipidemia** | **Denominator:** LDL $\geq 130$ (mg/dL)<br>2 readings $\geq 30$ days apart and no prior diagnosis.<br>**Numerator:** New diagnosis of any of the following:<br>ICD-9: 272.0, 272.2, 272.3, 272.4, 272.9<br>ICD-10: E78.0, E78.2, E78.3, E78.4, E78.5, E78.7, E78.8, E78.9 |

### Data Analysis and Statistical Approaches

Data extraction was done with custom PySpark scripts using Spark (v2.1.0). Preprocessing and summary statistics were performed using the pandas (v0.24.1) and NumPy (v1.16.2) Python libraries. Visualizations were produced with the Matplotlib (v3.0.3) and seaborn (v0.9.0) Python libraries. To model the time to diagnosis, we employed the Kaplan-Meier estimation method for survival analysis using the lifelines (v0.20.0) Python library. All study-specific scripts were reviewed by an independent analyst.

Patients with an existing (prior to first signal) or early (recorded between the first and second signal) diagnosis were excluded from the time to diagnosis analysis. The duration for the survival analysis (equivalent to “survival time”) was the number of days between the date of the second signal and the date of diagnosis. For those who were never diagnosed, it was defined as the number of days between the date of the second signal and the date of the most recent encounter, at which point they were censored due to lack of additional follow-up.

## Results

### Frequency of Clinical and Computed Diagnoses

To assess the temporal consistency of our data extract and ensure that there was no large change in coding practices for observations or diagnoses over time, we first assessed the frequency of extracted data elements. The number of diagnoses (Figure 1A) and observations (Figure 1B) over the study period remained qualitatively stable and no loss of observation or diagnostic codes was observed over time. The number of observations and diagnoses did increase over the study period as a unified EHR was brought online throughout the health system.

**Figure 1:**
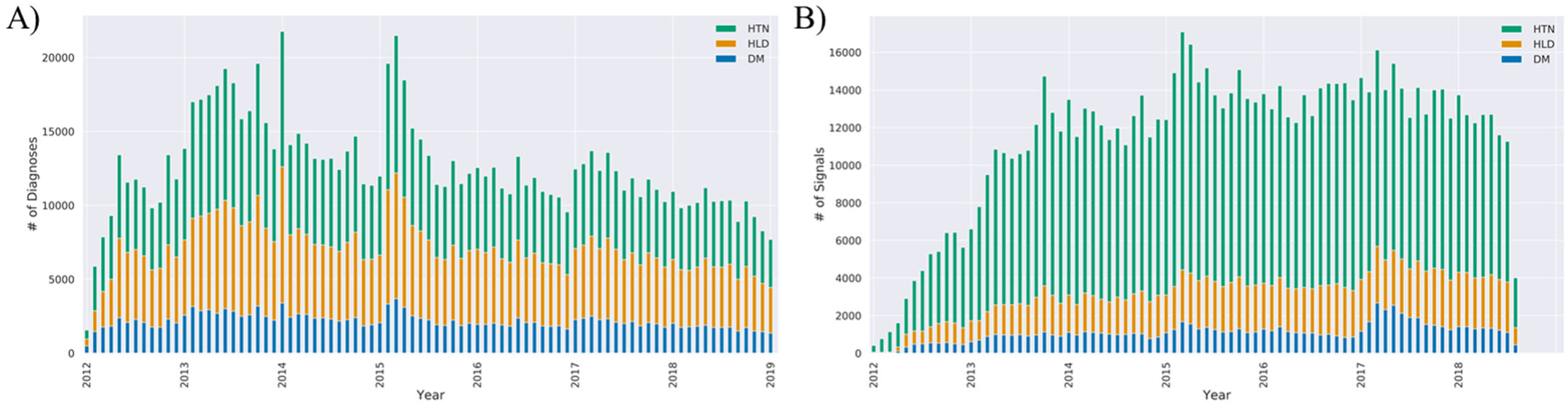
Frequency of structured EHR diagnoses over study period. Stacked bar charts showing the monthly counts of (A) structured diagnoses and (B) signals, or measurements above the threshold, over the study analysis period for hypertension (HTN), diabetes mellitus (DM), and hyperlipidemia (HLD).

We defined a signal of disease as an observation above the threshold. In our cohort, we computed a diagnosis of HTN in 245,711 patients, HLD in 45,098 patients, and DM in 33,153 patients who met our criteria for two signals separated by at least thirty days. Of these patients, a pre-existing, structured diagnosis of HTN, HLD, or DM, was present in the EHR for 42.0%, 37.4%, and 73.1% of patients, respectively, before the first signal was identified (Figure 2). For patients with a new diagnosis, there was a large degree of variability in the presence of structured diagnostic codes among the conditions we assessed. For DM, 89.8% of patients received an early structured diagnosis, meaning a structured diagnosis was recorded in the EHR in the window between the first and second signal. However, for those with HTN or HLD, only 36.4% and 51.3%, respectively, received an early clinical diagnosis. Similarly, 39.8% of those with computed HTN and 21.6% of those with a computed HLD diagnosis never received a structured diagnosis in the EHR, while only 1.0% of those with DM lacked a structured, clinical diagnosis.

**Figure 2:**
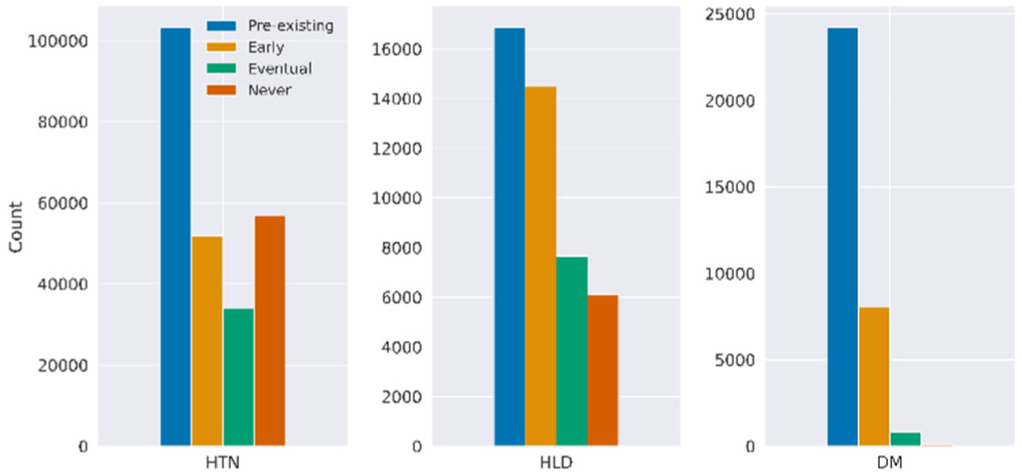
Frequency and temporality of structured diagnoses. For patients with a structured EHR diagnosis, the number of patients who had a pre-existing (preceded the first signal), early (recorded between the first and second signal), eventual (recorded after the computed diagnosis, or second signal), and the number of patients who never had a structured diagnosis recorded in the EHR.

### Origin of First Clinical Diagnosis

We also assessed the source of the first structured diagnosis to determine whether it came from a provider-entered, billing, or claims-based diagnosis. We found that the first source was similar among all three conditions, with most diagnoses being provider-entered within the medical history or problem list, followed by billing-related diagnoses (Figure 3). Only a small proportion of diagnoses were first identified via returned claims within our local data set.

**Figure 3:**
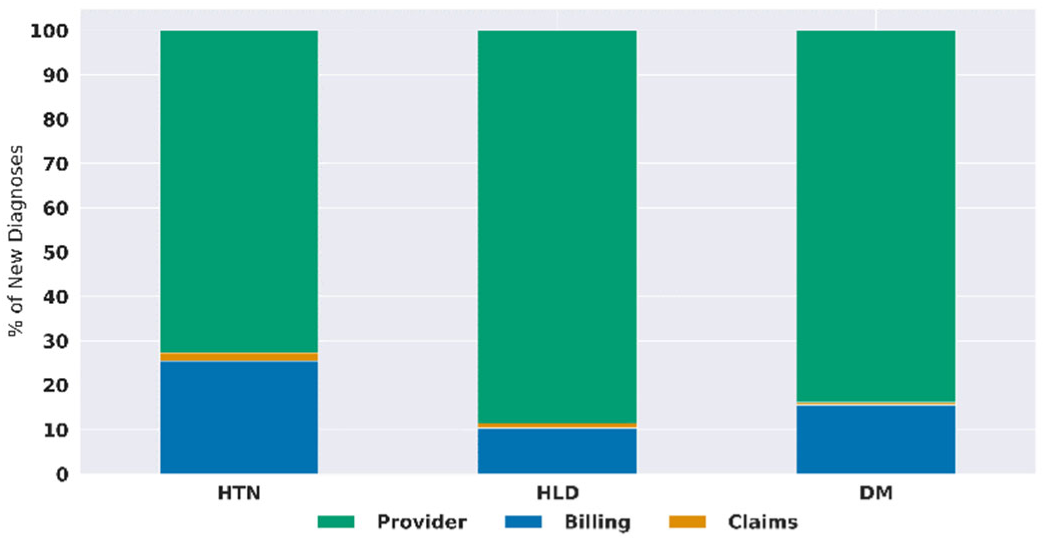
Source of initial diagnosis. The frequency of diagnosis source (provider-entered history or problem list, provider-entered billing, or returned claims) of the first structured diagnosis for patients with a new diagnosis of hypertension (HTN), diabetes mellitus (DM), or hyperlipidemia (HLD).

### Temporality of Clinical and Computed Diagnoses

Since the timing of diagnosis is relevant to cohort creation, we determined the delay in availability of the first structured diagnosis compared with when a diagnosis could be computed from other data available in the EHR. To define a consistent starting time, patients with a pre-existing diagnosis (present prior to first signal) or early diagnosis (occurring between the first and second signal) were excluded. Within this cohort, the mean time for a structured diagnosis to be recorded ranged from a minimum of approximately 100 days for DM to a maximum of nearly 600 days for HTN (Figure 4) from the time of the computed diagnosis. The temporal delay varied by the source of diagnosis, with provider-entered diagnoses having the shortest interval in all conditions and claims-based diagnoses having the longest. It should be noted that very few patients had a claim as the first occurrence of a structured diagnosis (n=633, 128, and 4 for HTN, HLD, and DM, respectively).

**Figure 4:**
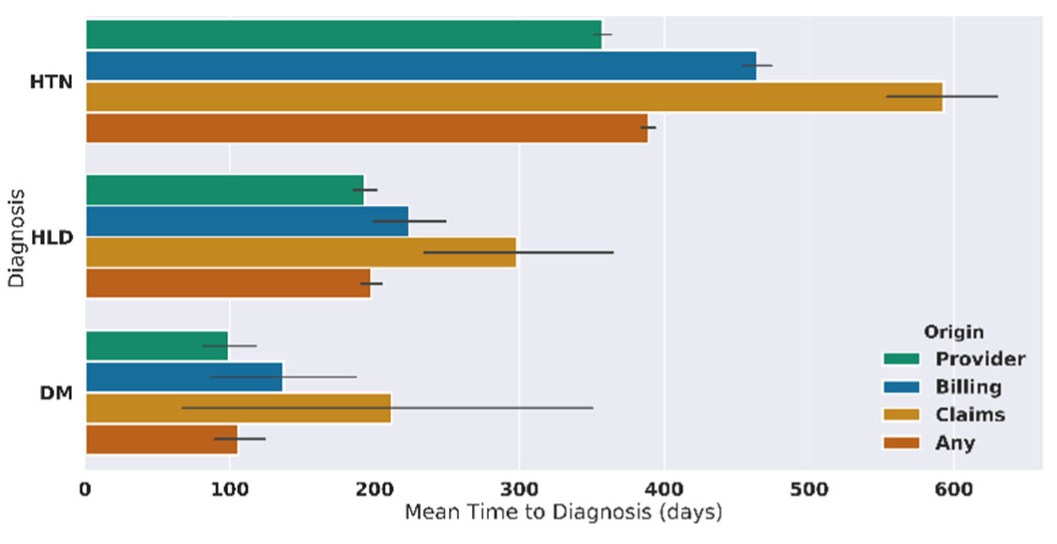
Mean time from computed to recorded diagnosis. The mean time in days to receive a structured diagnosis from the time of the second signal. Means were calculated using the number of days to initial diagnosis.

To assess the timeliness of structured diagnoses, we estimated the likelihood of having a structured diagnosis present in the EHR using the Kaplan-Meier estimation method, where the last encounter was used as the censor date if a discrete diagnosis was not found. The likelihood of not having a structured diagnosis also varied by condition (Figure 5). While those with DM had over a 90% chance of having a structured diagnosis recorded at 2 years, those with HLD had less than a 60% chance and those with HTN had less than a 40% chance of having a structured diagnosis present at the same timepoint. The mean time from the second signal of disease to censoring for those who did not receive a diagnosis was 726 days, 600 days, and 503 days for HTN, HLD, and DM, respectively.

**Figure 5:**
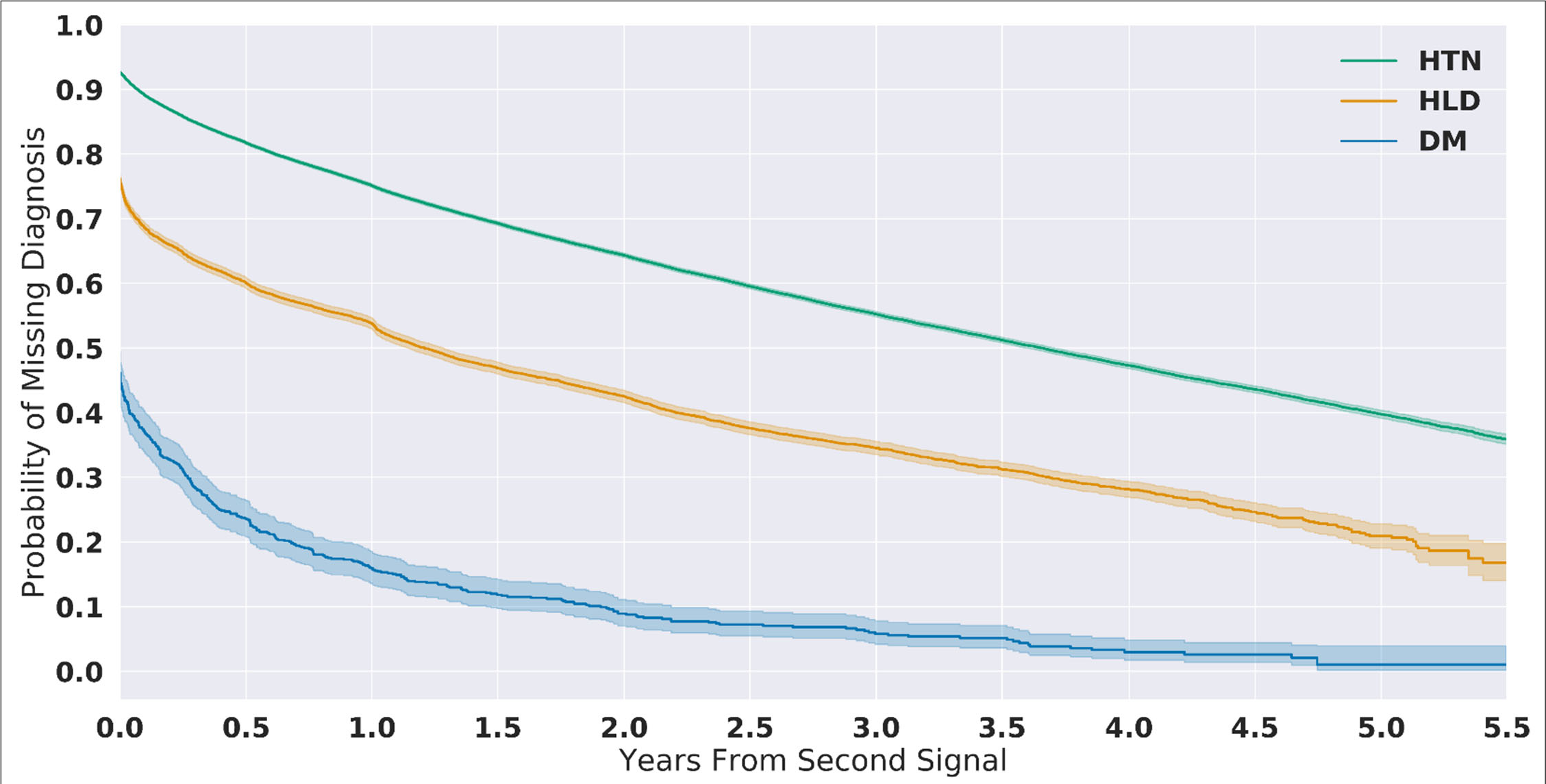
Temporal probability of the absence of a structured diagnosis. Kaplan-Meier estimated probability that a patient is missing a structured diagnosis after a computed diagnosis is made (presence of two signals) for patients with hypertension (HTN), diabetes mellitus (DM), or hyperlipidemia (HLD).

## Discussion

The primary finding of this study was that structured diagnoses within the EHR for HTN, HLD, and DM can have a marked delay in being recorded compared with the time a diagnosis can be computed from other EHR data. For three common diseases, the average time from which a diagnosis could be computed from laboratory values preceded the manual recording of a structured diagnosis by as much as 389 days. In addition, even one year after a diagnosis can be computed, a large percentage of patients do not have an equivalent structured diagnosis recorded in the EHR. Therefore, while the EHR has several potential advantages to other sources of RWD and can be accessed in near real-time from a technical perspective, the recording of clinical information within the structured history and problem list is less sensitive and delayed compared to identifying computed diagnoses. Thus, studies based on RWD, the approach to extracting information from the EHR may affect its quality.

The EHR contains a detailed record of a patient’s clinical history but extracting this history from the structured and unstructured fields that data can reside in remains a challenge. Our work extends the prior literature, which have focused on the completeness and accuracy of the problem list compared to manual adjudication or next generation phenotyping approaches.^22,23,25,30,37,38^ The delay in recording a structured diagnosis has the potential to impact the development of cohorts and outcome ascertainment from RWD because analyses based on structured diagnostic codes could preferentially identify patients with a longer history of disease. In addition, analyses limited to patients with more recent data are likely to have a lower prevalence of disease than cohorts with a longer history, which may bias historic comparisons with synthetic or external control arms. Finally, if data are obtained from multiple institutions with varying local diagnostic patterns, additional biases may be introduced to multi-site studies.

While concerns of EHR data completeness are often described as data collection and quality issues, this is, in many cases, primarily a concern when the data are used for secondary research purposes.^6,39,40^ When assessed from a clinical perspective, information related to a disorder, such as blood pressure measurements or documentation within an unstructured clinical note, can be used by a healthcare provider to draw equivalent conclusions, despite the high potential to be missed during automated digital extraction. Therefore, EHR data may not be missing or of low quality, but are rather collected for clinical, rather than research, purposes. Even with these limitations, EHR data can add significant value when analyzed appropriately. For example, others have demonstrated that what may often be described as noise within EHR data, such as frequency of measurements or presence of repeat diagnoses, can actually be used to predict patient outcome and the temporality of clinical conditions.^31,34,41^

Despite the potential concerns related to the use of EHR described here, it is important to acknowledge that similar issues can also be found in clinical research and manually adjudicated data sets, such as disease registries. Several studies have shown significant variability in the accuracy and inter-rater reliability of manual data abstraction. One case study by the Office of the Inspector General for the Department of Health and Human Services found that manual nurse review identified 78% (93 of 120) of adverse events in the study population.^42^ Similarly, patient report, a common source for clinical research studies, has been found to over-or under-represent even major healthcare events, such as readmission, in nearly 30% of cases. ^43^ Therefore, strategies to better understand and use RWD to augment data collected through traditional methods have the potential to increase the accuracy and completeness of patient history, clinical events, and healthcare outcomes.

This study has several limitations. First, data were collected from a single site within a healthcare system. However, our findings for the number of missing diagnoses for those with HTN were consistent with previously published studies.^26,27^ This work also focused on only three phenotypes which could be reliably identified from clinical measurements and laboratory testing, all of which were chronic diseases, and did not assess for more complex diagnoses. Finally, we did not assess the clinical impact of missing structured diagnoses,

While strategies to assess data quality and account for variations in data collection for clinical research data have been developed, access to and use of RWD remains a new and rapidly evolving field. Like diagnostic laboratory tests, methods to extract data from the EHR can be viewed as assays with varying sensitivity, specificity, and window periods. Work by the Electronic Medical Records and Genomics (eMERGE)^44^ and OHDSI^38,45^ networks, among others, to create standardized next generation phenotypes will continue to improve our ability to identify clinical cohorts and outcomes, but standardized strategies to assess whether RWD are fit-for-purpose and new methods to reduce bias in data collection will need ongoing, and likely use case-specific, assessment.

## Data Availability

As a large-scale quality analysis of clinical electronic health record data, raw data are not publicly available.

## Acknowledgements

We would like to acknowledge Aeron Small for his review of this manuscript. We would also like to acknowledge Charlie Torre, Jr, William Byron, and Nathaniel Price for their assistance with maintaining the underlying infrastructure for the data used in this analysis.

## Author Contributions

W.S. conceived the study and led the design and analysis. P.Y. carried out the data extraction and analyses. A.C. assisted with data analysis and independent code review. W.S., P.Y., A.C., B.M., Z.L., R.J., and H.K. discussed results and implications and all contributed to the writing and editing of the manuscript. All authors have approved the final manuscript.

## Competing Interests

Harlan Krumholz works under contract with the Centers for Medicare & Medicaid Services to support quality measurement programs; was a recipient of a research grant, through Yale, from Medtronic and the U.S. Food and Drug Administration to develop methods for post-market surveillance of medical devices; was a recipient of a research grant with Medtronic and is the recipient of a research grant from Johnson & Johnson, through Yale University, to support clinical trial data sharing; was a recipient of a research agreement, through Yale University, from the Shenzhen Center for Health Information for work to advance intelligent disease prevention and health promotion; collaborates with the National Center for Cardiovascular Diseases in Beijing; receives payment from the Arnold & Porter Law Firm for work related to the Sanofi clopidogrel litigation, from the Ben C. Martin Law Firm for work related to the Cook Celect IVC filter litigation, and from the Siegfried and Jensen Law Firm for work related to Vioxx litigation; chairs a Cardiac Scientific Advisory Board for UnitedHealth; was a participant/participant representative of the IBM Watson Health Life Sciences Board; is a member of the Advisory Board for Element Science, the Advisory Board for Facebook, and the Physician Advisory Board for Aetna; and is the co-founder of HugoHealth, a personal health information platform, and co-founder of Refactor Health, an AI-augmented data management platform for healthcare.

Wade Schulz was an investigator for a research agreement, through Yale University, from the Shenzhen Center for Health Information for work to advance intelligent disease prevention and health promotion; collaborates with the National Center for Cardiovascular Diseases in Beijing; is a technical consultant to HugoHealth, a personal health information platform, and co-founder of Refactor Health, an AI-augmented data management platform for healthcare; is a consultant for Interpace Diagnostics Group, a molecular diagnostics company.

The other co-authors report no potential competing interests.

## Notes

### Funding Statement

No external funding was received in support of this specific study.

## References

1. Mulder R, Singh AB, Hamilton A, et al. The limitations of using randomised controlled trials as a basis for developing treatment guidelines. Evid Based Ment Health 2018;21(1):4–6. doi:10.1136/eb-2017-102701.

2. Booth CM, Tannock IF. Randomised controlled trials and population-based observational research: partners in the evolution of medical evidence. Br. J. Cancer 2014;110(3):551–555. doi:10.1038/bjc.2013.725.

3. Sherman RE, Anderson SA, Dal Pan GJ, et al. Real-World Evidence - What Is It and What Can It Tell Us? N. Engl. J. Med. 2016;375(23):2293–2297. doi:10.1056/NEJMsb1609216.

4. Miksad RA, Abernethy AP. Harnessing the Power of Real-World Evidence (RWE): A Checklist to Ensure Regulatory-Grade Data Quality. Clin. Pharmacol. Ther. 2018;103(2):202–205. doi:10.1002/cpt.946.

5. Khosla S, White R, Medina J, et al. Real world evidence (RWE) – a disruptive innovation or the quiet evolution of medical evidence generation? [version 2; peer review: 2 approved]. F1000Res. 2018;7:111. doi:10.12688/f1000research.13585.2.

6. Hersh WR, Weiner MG, Embi PJ, et al. Caveats for the use of operational electronic health record data in comparative effectiveness research. Med. Care 2013;51(8 Suppl 3):S30–7. doi:10.1097/MLR.0b013e31829b1dbd.

7. Kim H-S, Kim JH. Proceed with Caution When Using Real World Data and Real World Evidence. J. Korean Med. Sci. 2019;34(4):e28. doi:10.3346/jkms.2019.34.e28.

8. Ryan PB, Madigan D, Stang PE, Overhage JM, Racoosin JA, Hartzema AG. Empirical assessment of methods for risk identification in healthcare data: results from the experiments of the Observational Medical Outcomes Partnership. Stat. Med. 2012;31(30):4401–4415. doi:10.1002/sim.5620.

9. Häyrinen K, Saranto K, Nykänen P. Definition, structure, content, use and impacts of electronic health records: a review of the research literature. Int. J. Med. Inform. 2008;77(5):291–304. doi:10.1016/j.ijmedinf.2007.09.001.

10. ICD - ICD-10-CM - International Classification of Diseases, Tenth Revision, Clinical Modification. Available at: https://www.cdc.gov/nchs/icd/icd10cm.htm. Accessed December 2, 2019.

11. Mitchell JB, Bubolz T, Paul JE, et al. Using Medicare claims for outcomes research. Med. Care 1994;32(7 Suppl):JS38–51.

12. Blumenthal S. The use of clinical registries in the united states: A landscape survey. EGEMS (Wash. DC) 2017;5(1):26. doi:10.5334/egems.248.

13. Birnbaum HG, Cremieux PY, Greenberg PE, LeLorier J, Ostrander JA, Venditti L. Using healthcare claims data for outcomes research and pharmacoeconomic analyses. Pharmacoeconomics 1999;16(1):1–8. doi:10.2165/00019053-199916010-00001.

14. Hoque DME, Kumari V, Hoque M, Ruseckaite R, Romero L, Evans SM. Impact of clinical registries on quality of patient care and clinical outcomes: A systematic review. PLoS One 2017;12(9):e0183667. doi:10.1371/journal.pone.0183667.

15. Mues KE, Liede A, Liu J, et al. Use of the Medicare database in epidemiologic and health services research: a valuable source of real-world evidence on the older and disabled populations in the US. Clin Epidemiol 2017;9:267–277. doi:10.2147/CLEP.S105613.

16. Krumholz HM, Lin Z, Drye EE, et al. An administrative claims measure suitable for profiling hospital performance based on 30-day all-cause readmission rates among patients with acute myocardial infarction. Circ Cardiovasc Qual Outcomes 2011;4(2):243–252. doi:10.1161/CIRCOUTCOMES.110.957498.

17. Ferver K, Burton B, Jesilow P. The Use of Claims Data in Healthcare Research. Open Public Health J. 2009;2(1):11–24. doi:10.2174/1874944500902010011.

18. The PCORI Methodology Report | PCORI. Available at: https://www.pcori.org/research-results/about-our-research/research-methodology/pcori-methodology-report. Accessed December 3, 2019.

19. Jollis JG, Ancukiewicz M, DeLong ER, Pryor DB, Muhlbaier LH, Mark DB. Discordance of databases designed for claims payment versus clinical information systems. Implications for outcomes research. Ann. Intern. Med. 1993;119(8):844–850. doi:10.7326/0003-4819-119-8-199310150-00011.

20. Hartzema AG, Racoosin JA, MaCurdy TE, Gibbs JM, Kelman JA. Utilizing Medicare claims data for real-time drug safety evaluations:is it feasible? Pharmacoepidemiol Drug Saf 2011;20(7):684–688. doi:10.1002/pds.2143.

21. Subramanian S, Tangka FKL, Beebe MC, Trebino D, Weir HK, Babcock F. The cost of cancer registry operations: Impact of volume on cost per case for core and enhanced registry activities. Eval. Program Plann. 2016;55:1–8. doi:10.1016/j.evalprogplan.2015.11.005.

22. Wright A, McCoy AB, Hickman T-TT, et al. Problem list completeness in electronic health records: A multi-site study and assessment of success factors. Int. J. Med. Inform. 2015;84(10):784–790. doi:10.1016/j.ijmedinf.2015.06.011.

23. Singer A, Kroeker AL, Yakubovich S, Duarte R, Dufault B, Katz A. Data quality in electronic medical records in Manitoba: Do problem lists reflect chronic disease as defined by prescriptions? Can Fam Physician 2017;63(5):382–389.

24. Holmes C, Brown M, Hilaire DS, Wright A. Healthcare provider attitudes towards the problem list in an electronic health record: a mixed-methods qualitative study. BMC Med Inform Decis Mak 2012;12:127. doi:10.1186/1472-6947-12-127.

25. Szeto HC, Coleman RK, Gholami P, Hoffman BB, Goldstein MK. Accuracy of computerized outpatient diagnoses in a Veterans Affairs general medicine clinic. Am. J. Manag. Care 2002;8(1):37–43.

26. Smith SM, McAuliffe K, Hall JM, et al. Hypertension in florida: data from the oneflorida clinical data research network. Prev Chronic Dis 2018;15:E27. doi:10.5888/pcd15.170332.

27. Banerjee D, Chung S, Wong EC, Wang EJ, Stafford RS, Palaniappan LP. Underdiagnosis of hypertension using electronic health records. Am. J. Hypertens. 2012;25(1):97–102. doi:10.1038/ajh.2011.179.

28. Hripcsak G, Ryan PB, Duke JD, et al. Characterizing treatment pathways at scale using the OHDSI network. Proc. Natl. Acad. Sci. USA 2016;113(27):7329–7336. doi:10.1073/pnas.1510502113.

29. Toh S, Rasmussen-Torvik LJ, Harmata EE, et al. The National Patient-Centered Clinical Research Network (PCORnet) Bariatric Study Cohort: Rationale, Methods, and Baseline Characteristics. JMIR Res Protoc 2017;6(12):e222. doi:10.2196/resprot.8323.

30. Hripcsak G, Albers DJ. Next-generation phenotyping of electronic health records. J. Am. Med. Inform. Assoc. 2013;20(1):117–121. doi:10.1136/amiajnl-2012-001145.

31. Perotte A, Hripcsak G. Temporal properties of diagnosis code time series in aggregate. IEEE J Biomed Health Inform 2013;17(2):477–483. doi:10.1109/JBHI.2013.2244610.

32. Elliott AF, Davidson A, Lum F, et al. Use of electronic health records and administrative data for public health surveillance of eye health and vision-related conditions in the United States. Am. J. Ophthalmol. 2012;154(6 Suppl):S63–70. doi:10.1016/j.ajo.2011.10.002.

33. White Paper: The Benefit of Using Both Claims Data and Electronic Medical Record Data in Health Care Analysis. Optum, Inc.; 2014. Available at: https://www.optum.com/resources/library/benefit-using-both-claims-data-electronic-medical-record-data-health-care-analysis.html. Accessed August 2, 2019.

34. Agniel D, Kohane IS, Weber GM. Biases in electronic health record data due to processes within the healthcare system: retrospective observational study. BMJ 2018;361:k1479. doi:10.1136/bmj.k1479.

35. McPadden J, Durant TJ, Bunch DR, et al. Health care and precision medicine research: analysis of a scalable data science platform. J. Med. Internet Res. 2019;21(4):e13043. doi:10.2196/13043.

36. PCORnet. PCORnet Common Data Model (CDM). PCORnet Common Data Model (CDM) 2017. Available at: http://www.ohdsi.org/web/wiki/doku.php?id=documentation:cdm:single-page. Accessed August 11, 2018.

37. Blecker S, Katz SD, Horwitz LI, et al. Comparison of approaches for heart failure case identification from electronic health record data. JAMA Cardiol. 2016;1(9):1014–1020. doi:10.1001/jamacardio.2016.3236.

38. Hripcsak G, Shang N, Peissig PL, et al. Facilitating phenotype transfer using a common data model. J. Biomed. Inform. 2019:103253. doi:10.1016/j.jbi.2019.103253.

39. Weiskopf NG, Weng C. Methods and dimensions of electronic health record data quality assessment: enabling reuse for clinical research. J. Am. Med. Inform. Assoc. 2013;20(1):144–151. doi:10.1136/amiajnl-2011-000681.

40. Weiner MG, Embi PJ. Toward reuse of clinical data for research and quality improvement: the end of the beginning? Ann. Intern. Med. 2009;151(5):359–360.

41. Hripcsak G, Albers DJ, Perotte A. Exploiting time in electronic health record correlations. J. Am. Med. Inform. Assoc. 2011;18 Suppl 1:i109–15. doi:10.1136/amiajnl-2011-000463.

42. Golladay KK, Collins AB, Ashcraft A, et al. Adverse Events in Hospitals: Methods for Identifying Events. Department of Health and Human Services; 2010:60.

43. Krishnamoorthy A, Peterson ED, Knight JD, et al. How Reliable are Patient-Reported Rehospitalizations? Implications for the Design of Future Practical Clinical Studies. J. Am. Heart Assoc. 2016;5(1). doi:10.1161/JAHA.115.002695.

44. Electronic Medical Records and Genomics (emerge) Network. Available at: https://emerge.mc.vanderbilt.edu/about-emerge/. Accessed October 31, 2018.

45. Hripcsak G, Duke JD, Shah NH, et al. Observational health data sciences and informatics (OHDSI): opportunities for observational researchers. Stud. Health Technol. Inform. 2015;216:574–578.

